# Identifying Ashkenazi Jewish *BRCA1/2* founder variants in individuals who do not self-report Jewish ancestry

**DOI:** 10.1101/19001032

**Authors:** Ruth I. Tennen, Sarah B. Laskey, Bertram L. Koelsch, Matthew H. McIntyre, The 23andMe Health Team, Joyce Y. Tung

**Affiliations:** 23andMe, Inc., 899 W Evelyn Ave, Mountain View, CA, 94041 USA

**Author notes:** Correspondence should be addressed to J.Y.T.

**Keywords:** BRCA1, BRCA2, genetic testing, Ashkenazi Jewish, family history

## Abstract

Current guidelines recommend *BRCA1* and *BRCA2* genetic testing for individuals with a personal or family history of certain cancers. Three *BRCA1/2* founder variants — 185delAG (c.68_69delAG), 5382insC (c.5266dupC), and 6174delT (c.5946delT) — are common in the Ashkenazi Jewish population. We characterized a cohort of more than 2,800 research participants in the 23andMe database who carry one or more of the three Ashkenazi Jewish founder variants, evaluating two characteristics that are typically used to recommend individuals for *BRCA* testing: self-reported Jewish ancestry and family history of breast, ovarian, prostate, or pancreatic cancer. Of the 1,967 carriers who provided self-reported ancestry information, 21% did not self-report any Jewish ancestry; of these individuals, more than half (62%) do have detectable Ashkenazi Jewish genetic ancestry. In addition, of the 343 carriers who provided both ancestry and family history information, 44% did not have a first-degree family history of a *BRCA*-related cancer and, in the absence of a personal history of cancer, would therefore be unlikely to qualify for clinical genetic testing. These findings provide support for the growing call for broader access to *BRCA* genetic testing.

## BACKGROUND

Pathogenic variants in the *BRCA1* and *BRCA2* genes are linked to an increased risk for female breast and ovarian cancer (including early-onset breast cancer), male breast cancer, prostate cancer, pancreatic cancer, and certain other cancers [1]. These variants are highly penetrant: Women with a variant have a 45-85% chance of developing breast cancer and up to a 46% chance of developing ovarian cancer by age 70 [2]. However, increased surveillance and prophylactic surgery (mastectomy and salpingo-oophorectomy) can greatly reduce the risk of breast and ovarian cancer in women carrying a *BRCA1* or *BRCA2* mutation [3].

The prevalence of pathogenic *BRCA1* and *BRCA2* variants is estimated to be between 1 in 300 and 1 in 800 in the general population [1,2]. Among individuals of Ashkenazi Jewish descent, three *BRCA1/2* founder variants — 185delAG (c.68_69delAG), 5382insC (c.5266dupC), and 6174delT (c.5946delT) — are present at a frequency of ∼1 in 40 [1].

Current U.S. guidelines limit *BRCA1*/2 genetic testing to individuals with a personal or family history of a relevant cancer, including early-onset breast cancer, multiple primary breast cancers, ovarian cancer, and certain other cancers [1,2,4]. In addition, Ashkenazi Jewish ancestry is sometimes used to recommend screening for individuals with a personal or family history of a single breast cancer at any age [1]. However, recent studies have found that about 50% of *BRCA* carriers have little or no family history of a relevant cancer [5-8]. These individuals would likely not qualify for clinical genetic testing unless they developed cancer themselves, representing a missed opportunity for cancer prevention. Because *BRCA* variants predispose to very high breast and ovarian cancer risks even among carriers without a family history [5,9], these findings have spurred calls for broader access to *BRCA* genetic testing, among Ashkenazi Jews and in the general population [5-7,10,11].

The 23andMe database provides an ethnically diverse, generally unselected group of genotyped individuals. We sought to characterize a cohort of individuals who carry one or more of the three Ashkenazi Jewish founder variants as related to two characteristics that are typically used to recommend individuals for *BRCA* testing: self-reported Jewish ancestry and family history of breast, ovarian, prostate, or pancreatic cancer. We focused on these two characteristics because they can enable individuals to learn their *BRCA* status before developing cancer, thus providing opportunities for cancer prevention and/or early detection.

## METHODS

Consented participants were drawn from the customer base of 23andMe. Data on Jewish ancestry and family cancer history were collected by self-report via online surveys. See Declarations for informed consent and protocol details; see Supplementary Information for survey questions. Analyses were run on phenotypic data collected before October 10, 2017.

DNA extraction and genotyping were performed on saliva samples by CLIA-certified and CAP-accredited clinical laboratories of Laboratory Corporation of America. Samples were genotyped on one of four custom Illumina genotyping arrays: the HumanHap550+ Bead chip (two versions), the OmniExpress+ Bead chip, or a fully custom array; see [12] for additional details. Proportions of Ashkenazi Jewish genetic ancestry were estimated via an analysis of local genetic ancestry as described in [13]. To account for imprecision in genetic ancestry estimates, we characterized estimates of Ashkenazi Jewish genetic ancestry <1% as “not detectable.”

## RESULTS

We identified 2,853 individuals who carry one or more of the three Ashkenazi Jewish *BRCA1/2* founder variants (Table 1).

**Table 1.** Demographics of individuals carrying one or more of the three Ashkenazi Jewish *BRCA* founder variants

| Age (years) | Men | Women |
| --- | --- | --- |
| Overall | 1539 | 1314 |
| 18-30 | 205 (13.3%) | 186 (14.2%) |
| 31-50 | 517 (33.6%) | 461 (35.1%) |
| 51-70 | 550 (35.7%) | 510 (38.8%) |
| 71+ | 267 (17.3%) | 157 (11.9%) |

We first characterized the ethnic backgrounds of the carriers. The three variants in this study are most common in people of Ashkenazi Jewish descent. However, among carriers who provided self-reported ancestry information, 21% did not report Jewish ancestry (Table 2).

**Table 2.** Self-reported Jewish ancestry in the cohort of *BRCA* carriers

|  |  |
| --- | --- |
| Total *BRCA* carriers | 2853 |
| Provided self-reported ancestry | 1967 (68.9%, of 2853) |
| - *BRCA1* 185delAG | 748 (38.0%, of 1967) |
| - *BRCA1* 5382insC | 415 (21.1%, of 1967) |
| - *BRCA2* 6174delT | 811 (41.2%, of 1967) |
| - Self-reported Jewish ancestry | 1552 (78.9%, of 1967) |
| - Did not self-report Jewish ancestry | 415 (21.1%, of 1967) |

The total number of *BRCA* variants detected exceeds the total number of individuals in the cohort because a small number of participants carry both a *BRCA1* and a *BRCA2* variant.

One possible explanation for the large fraction of carriers who did not report Jewish ancestry is that they were unaware of their Ashkenazi Jewish ancestry. To test this hypothesis, we explored the relationship between self-reported ancestry and genetic ancestry. As expected, individuals with a greater proportion of estimated Ashkenazi Jewish genetic ancestry were more likely to report Jewish ancestry (Table 3 and Figure 1); fewer than half of individuals with less than 20% Ashkenazi Jewish genetic ancestry (roughly equivalent to one grandparent or great-grandparent who was Ashkenazi Jewish) reported Jewish ancestry. Furthermore, most (62%, 258 of 415) of the *BRCA* carriers who reported no Jewish ancestry did have at least 1% Ashkenazi Jewish genetic ancestry. However, a lack of knowledge of Ashkenazi Jewish ancestry could not fully account for the 21% of carriers who did not report Jewish ancestry, as 8.4% (166 of 1,967) of individuals carrying an Ashkenazi Jewish founder variant had no detectable Ashkenazi Jewish genetic ancestry. This is consistent with the finding that at least two of these variants are also found in people of other ethnicities [14]. Nine individuals reported Jewish ancestry but had no detectable Ashkenazi Jewish genetic ancestry.

**Table 3.**
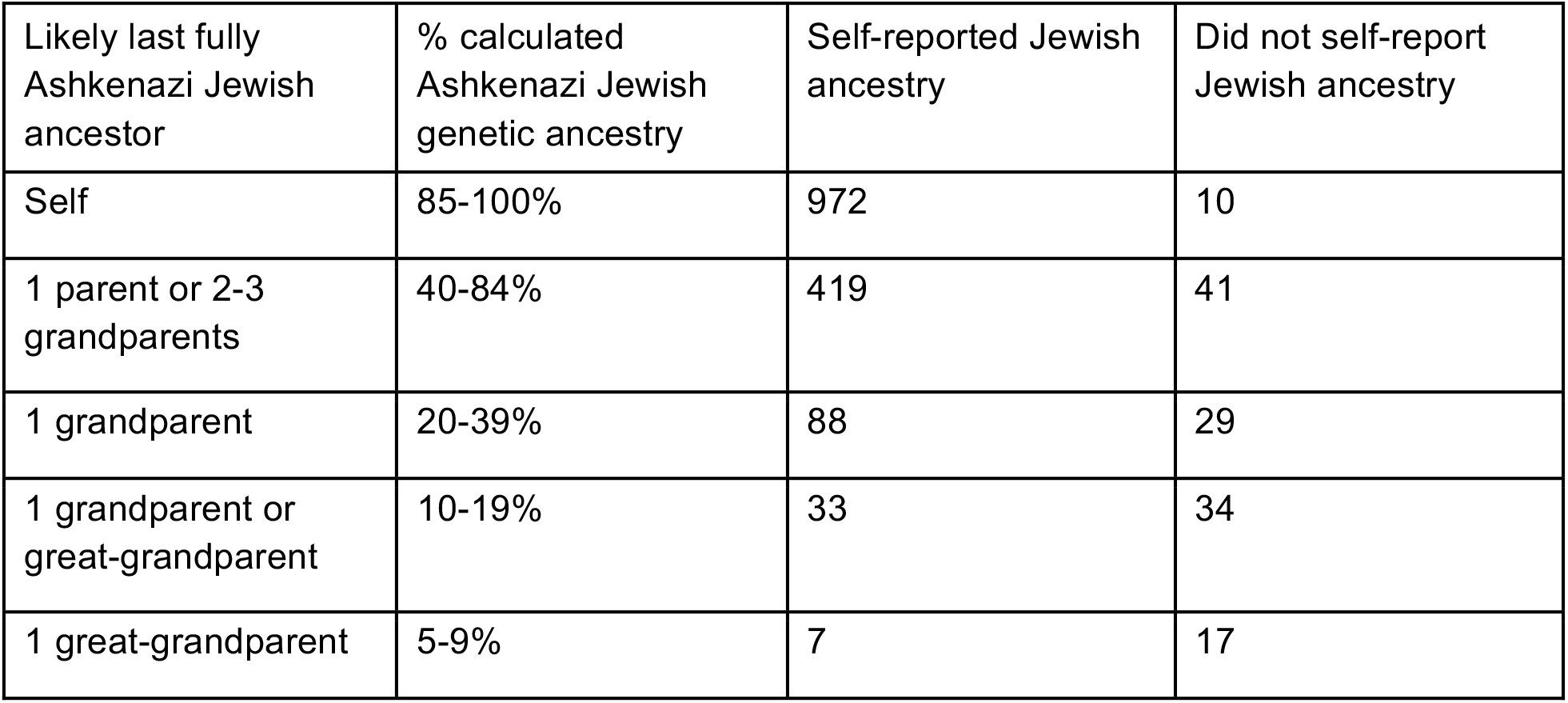
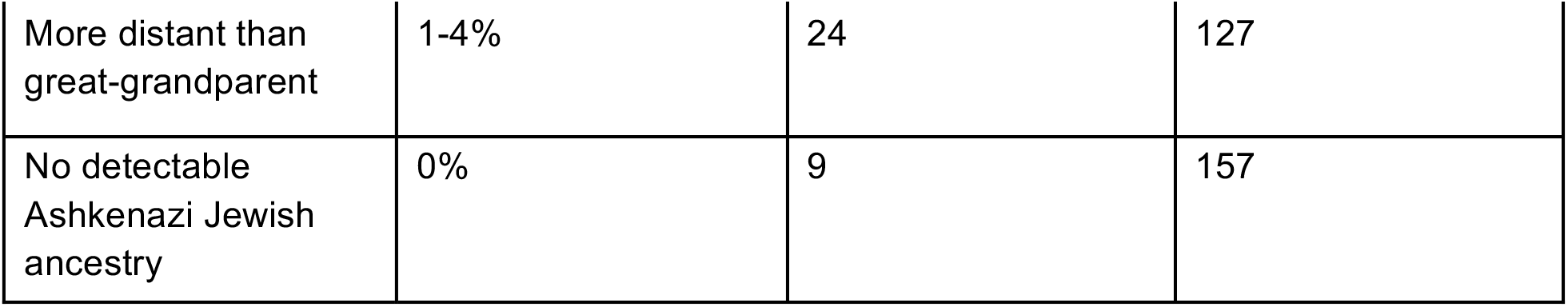
Self-reported Jewish ancestry vs. estimated Ashkenazi Jewish genetic ancestry in 1,967 *BRCA* carriers

**Figure 1.**
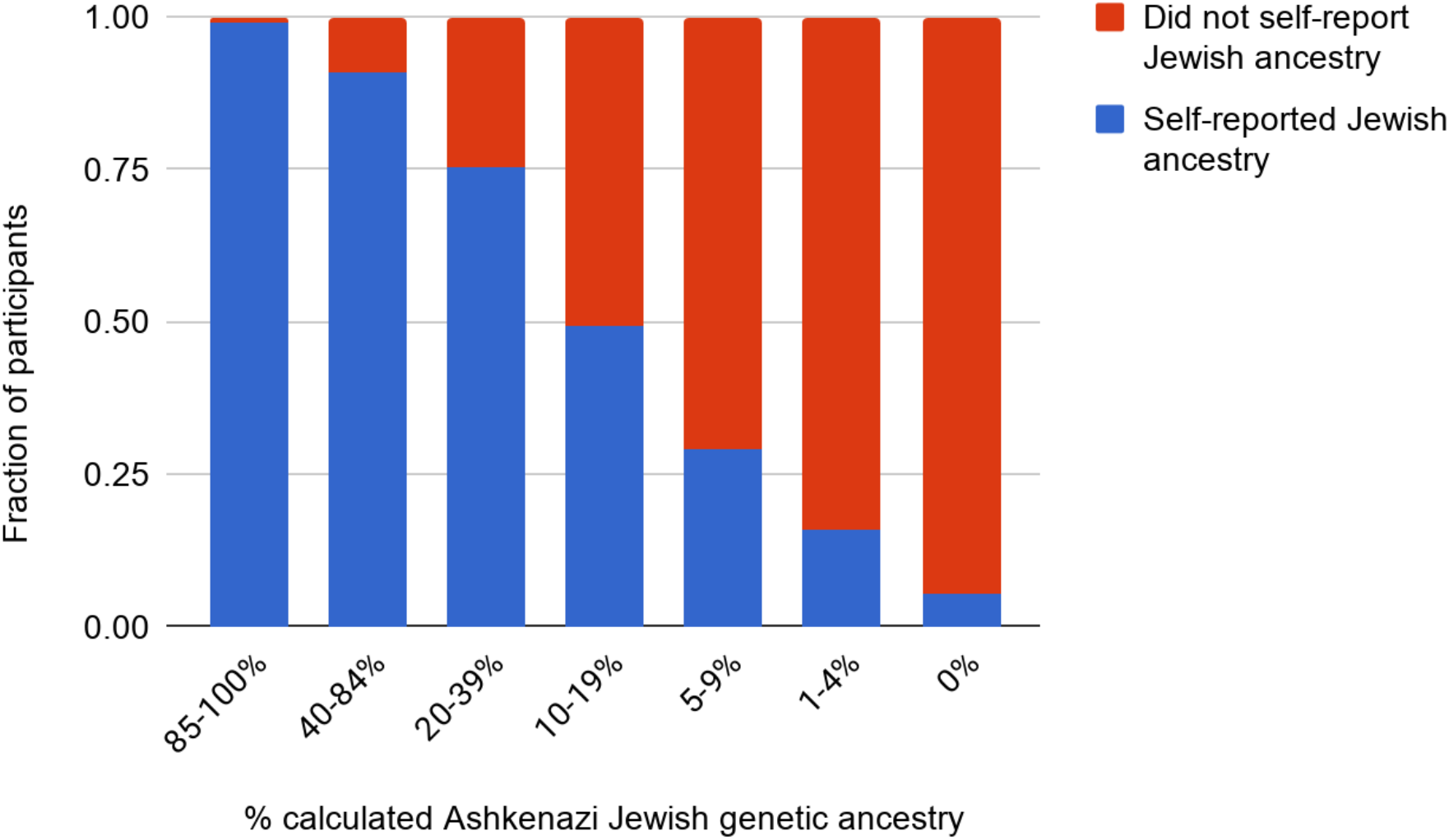
Self-reported Jewish ancestry vs. estimated Ashkenazi Jewish genetic ancestry in 1,967 *BRCA* carriers

The primary criterion for *BRCA* genetic testing is a personal or family history of breast, ovarian, or certain other cancers (including prostate and pancreatic cancer). We therefore assessed whether the carriers in our cohort had any family history of cancer. 393 carriers provided family history information.

Among participants who reported Jewish ancestry, 41% reported no first-degree family history of a *BRCA*-related cancer (Table 4). Similarly, 54% of participants who did not report Jewish ancestry reported no first-degree family history of cancer. Although our family history data differs substantially from guidelines used to determine genetic testing eligibility (which often include age of diagnosis and more than one cancer of certain types), this number is consistent with previous reports that about 50% of *BRCA* carriers would not be eligible for genetic testing based on family history alone [5-8].

**Table 4.**
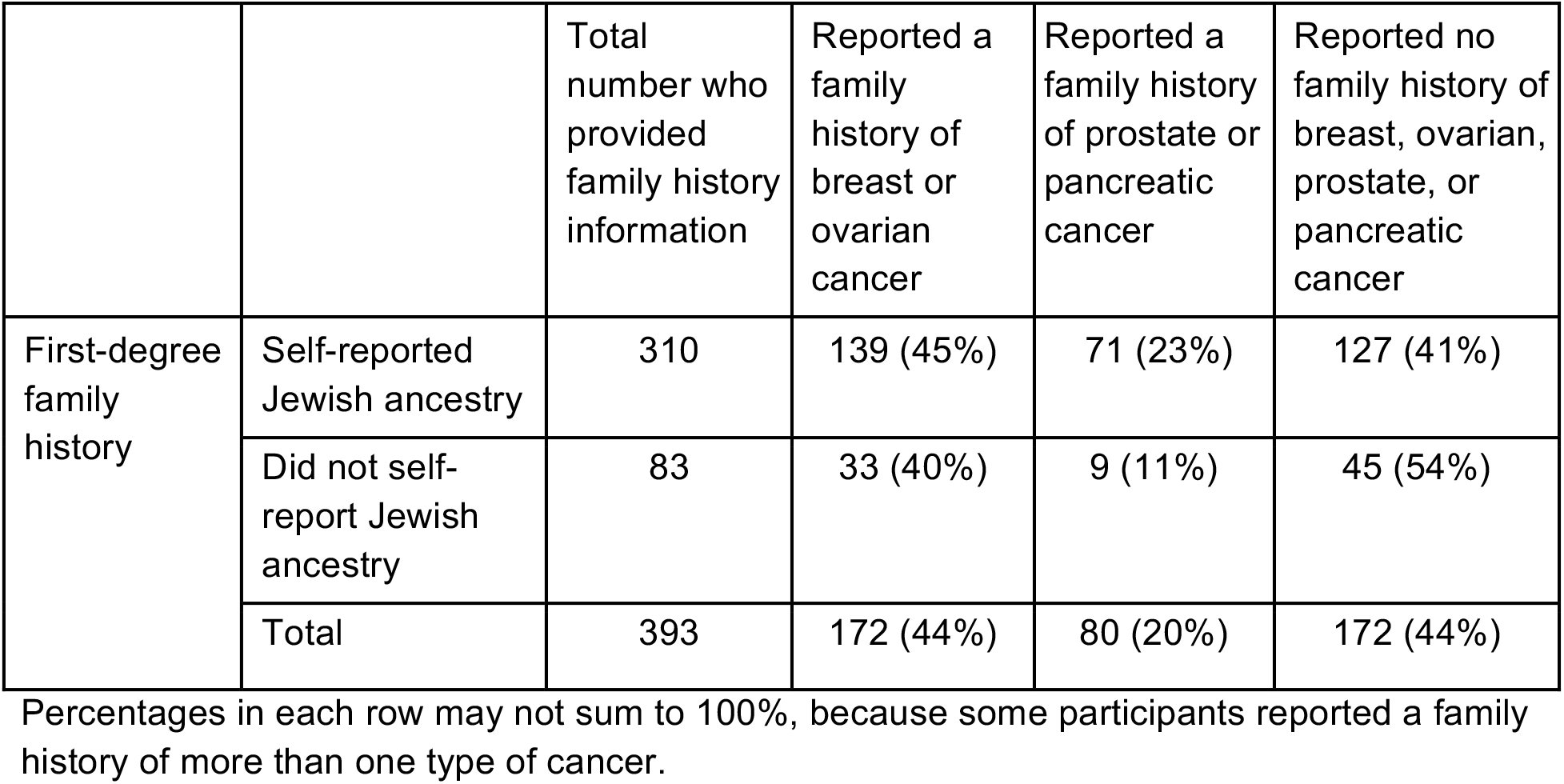
Self-reported family history of cancer in 393 *BRCA* carriers

## DISCUSSION

In this study, we describe a largely unselected cohort of ∼2,800 individuals who carry one or more of the three Ashkenazi Jewish *BRCA1/2* founder variants. In characterizing the ancestry and family cancer histories of these individuals, we made two key observations.

First, we found that a sizeable proportion (21%) of the carriers in our cohort do not self-report Jewish ancestry. Of these individuals, more than half (62%) do have detectable Ashkenazi Jewish genetic ancestry, although frequently in very low percentages (Table 3 and Figure 1). Interestingly, 8% of all carriers have no detectable Ashkenazi Jewish genetic ancestry, consistent with reports that *BRCA1* 185delAG and 5382insC are found in women of other ethnicities who are referred for clinical genetic testing [14].

Second, we observed that nearly half of individuals carrying an Ashkenazi Jewish *BRCA* variant have no first-degree family history of a *BRCA*-related cancer and, in the absence of a personal cancer history, would therefore be unlikely to qualify for clinical genetic testing. This percentage is consistent with published reports that about 50% of *BRCA* carriers lack a strong family history of cancer [5-8].

Limitations of this study include a potential ascertainment bias related to family cancer history, as individuals with such histories may be more likely to answer questions about family cancer history. In addition, due to the limited depth of our family history survey, we defined family history as having a first-degree relative with breast, ovarian, prostate, or pancreatic cancer; clinical testing criteria are typically stricter, requiring an early age of diagnosis and/or more than one affected family member. Together, these two points suggest that our estimate of the fraction of individuals who would be ineligible for testing based on family history alone under existing screening guidelines is likely lower than the true fraction. Finally, because not all individuals in this study provided ancestry and family history information, the number of individuals included in some analyses is fairly small.

Our data suggest that a sizeable fraction of individuals with detectable Ashkenazi Jewish genetic ancestry are unaware of that ancestry. This phenomenon is likely not unique to Ashkenazi Jewish ancestry. In addition to *BRCA*-related cancers, many other conditions are more common in specific ancestral groups, including Tay-Sachs disease, Canavan disease, and Gaucher disease type 1 in Ashkenazi Jews; sickle cell anemia in individuals with African ancestry; and beta-thalassemia in individuals with Mediterranean and certain other ancestries.

For individuals who are unaware of their genetic ancestry, perceived risk for diseases could thus differ substantially from actual risk, which could lead to missed opportunities for genetic screening, prevention, and early intervention.

In recent years, several groups have called for broader access to *BRCA* genetic testing among Ashkenazi Jews and among women in the general population [5-7,10,11]. Among Ashkenazi Jews, where testing for the three founder variants can identify most *BRCA* carriers, population-wide screening is cost-effective or even cost-saving [15]; in other ethnicities, more comprehensive genetic testing would be required to identify most individuals carrying a *BRCA* variant, but depending on the source of testing, this may also be cost-effective [16]. Our data provide support for this growing call for expanded *BRCA* testing. Such testing would enable women and men with a *BRCA* variant to learn their status, take steps to reduce their cancer risk, and encourage cascade testing of close family members.

## Data Availability

All data generated or analyzed during this study are included in the manuscript.

## DECLARATIONS

### Ethics approval and consent to participate

All participants provided informed consent and answered surveys online according to a research protocol approved by Ethical and Independent Review Services, an external AAHRPP-accredited institutional review board.

### Availability of data

All data generated or analyzed during this study are included in the manuscript.

### Authors’ contributions

Study concept and design: J.Y.T. and R.I.T. Acquisition and analysis of data: J.Y.T., S.B.L., B.L.K, and M.H.M. Interpretation of data: R.I.T., S.B.L., B.L.K, M.H.M., and J.Y.T. Drafting of the manuscript: R.I.T. Critical revision of the manuscript for important intellectual content: R.I.T., S.B.L., B.L.K, M.H.M., and J.Y.T. Support and infrastructure to enable the research presented here: The 23andMe Health Team.

## Acknowledgements

We thank the research participants and employees of 23andMe for making this work possible. We also thank Uta Francke, Jennifer C. McCreight, and Elizabeth S. Noblin for critical review of the manuscript; David A. Hinds for scientific input; and Catherine Wilson for supporting the ancestry and cancer family history surveys.

The 23andMe Health Team: Robert K. Bell, Katarzyna Bryc, Alison L. Chubb, Stacey B. Detweiler, Anne E. Greb, Esther Kim, Michaela Johnson, Joanna L. Mountain, Jamaica R. Perry, Jeffrey D. Pollard, Shirley Wu.

## SUPPLEMENTARY INFORMATION

### Jewish ancestry

Do you have any Jewish ancestry?

( ) Yes
( ) No
( ) I’m not sure

Participants who answered “Yes” were counted as self-reporting Jewish ancestry; participants who answered “No” were counted as not self-reporting Jewish ancestry; and participants who answered “I’m not sure” were not counted.

### Family history

Participants provided information about family history of cancer in one of three surveys, which included slightly different versions of the family history questions. Functionally, there were two versions of the family history questions (described below). Version 1 was in one survey while Version 2 was in two surveys. Relevant questions from each survey are excerpted below.

#### Version 1

Is your family’s history of cancer completely unknown to you for some reason, such as being adopted?

( ) Yes
( ) No

*Please indicate below whether your biological parents or grandparents have ever had cancer. [Grandparental history was not used*.]

Your mother

( ) Yes
( ) No
( ) I’m not sure

Your father

( ) Yes
( ) No
( ) I’m not sure

*Have any of your other biological relatives, listed below, had cancer?*

Your child(ren)

( ) Yes
( ) No
( ) Not applicable
( ) I’m not sure

Your sister(s)

( ) Yes
( ) No
( ) Not applicable
( ) I’m not sure

Your brother(s)

( ) Yes
( ) No
( ) Not applicable
( ) I’m not sure

{Where <female relative> could be “mother” or “sister”}

What type(s) of cancer was your <female relative> diagnosed with? Please check all that apply.

[ ] Adrenal gland cancer
[ ] Anal cancer
[ ] Bladder cancer
[ ] Biliary tract cancer
[ ] Brain cancer
[ ] Breast cancer
[ ] Cervical cancer
[ ] Colon/colorectal cancer
[ ] Endometrial or uterine cancer
[ ] Esophageal cancer
[ ] Gallbladder cancer
[ ] Hodgkin’s lymphoma
[ ] Kidney/renal cancer
[ ] Leukemia, any type
[ ] Liver/bile ducts (hepatobiliary) cancer
[ ] Lung cancer
[ ] Melanoma
[ ] Mouth (oral) cancer
[ ] Non-Hodgkin’s lymphoma
[ ] Pancreatic cancer
[ ] Ovarian cancer
[ ] Salivary gland cancer
[ ] Sarcoma, any type
[ ] Skin cancer (not melanoma)
[ ] Small intestine/duodenal cancer
[ ] Stomach/gastric cancer
[ ] Throat cancer
[ ] Thyroid cancer
[ ] Ureter/renal pelvis cancer
[ ] Vaginal cancer
[ ] Vulval cancer
[ ] Another type of cancer: _____
[ ] I’m not sure

{Where <male relative> could be “father” or “brother”}

What type(s) of cancer was your <male relative> diagnosed with? Please check all that apply.

[ ] Adrenal gland cancer
[ ] Anal cancer
[ ] Bladder cancer
[ ] Biliary tract cancer
[ ] Brain cancer
[ ] Breast cancer
[ ] Colon/colorectal cancer
[ ] Esophageal cancer
[ ] Gallbladder cancer
[ ] Hodgkin’s lymphoma
[ ] Kidney/renal cancer
[ ] Leukemia, any type
[ ] Liver/bile ducts (hepatobiliary) cancer
[ ] Lung cancer
[ ] Melanoma
[ ] Mouth (oral) cancer
[ ] Non-Hodgkin’s lymphoma
[ ] Pancreatic cancer
[ ] Prostate cancer
[ ] Salivary gland cancer
[ ] Sarcoma, any type
[ ] Skin cancer (not melanoma)
[ ] Small intestine/duodenal cancer
[ ] Stomach/gastric cancer
[ ] Testicular cancer
[ ] Throat cancer
[ ] Thyroid cancer
[ ] Ureter/renal pelvis cancer
[ ] Another type of cancer: _____
[ ] I’m not sure

What type(s) of cancer was your child(ren) diagnosed with? Please check all that apply.

[ ] Adrenal gland cancer
[ ] Anal cancer
[ ] Bladder cancer
[ ] Biliary tract cancer
[ ] Brain cancer
[ ] Breast cancer
[ ] Cervical cancer
[ ] Colon/colorectal cancer
[ ] Endometrial or uterine cancer
[ ] Esophageal cancer
[ ] Gallbladder cancer
[ ] Hodgkin’s lymphoma
[ ] Kidney/renal cancer
[ ] Leukemia, any type
[ ] Liver/bile ducts (hepatobiliary) cancer
[ ] Lung cancer
[ ] Melanoma
[ ] Mouth (oral) cancer
[ ] Non-Hodgkin’s lymphoma
[ ] Pancreatic cancer
[ ] Prostate cancer
[ ] Ovarian cancer
[ ] Salivary gland cancer
[ ] Sarcoma, any type
[ ] Skin cancer (not melanoma)
[ ] Small intestine/duodenal cancer
[ ] Stomach/gastric cancer
[ ] Testicular cancer
[ ] Throat cancer
[ ] Thyroid cancer
[ ] Ureter/renal pelvis cancer
[ ] Vaginal cancer
[ ] Vulval cancer
[ ] Another type of cancer: _____
[ ] I’m not sure

Have you ever been diagnosed with any of these hereditary cancer syndromes?

[ ] Cowden syndrome
[ ] Familial adenomatous polyposis
[ ] Hereditary breast and ovarian cancer syndrome (HBOC)
[ ] Li-Fraumeni syndrome
[ ] Lynch syndrome (hereditary non-polyposis colorectal cancer syndrome)
[ ] Multiple endocrine neoplasias
[ ] Von Hippel-Lindau Disease
[ ] I’m not sure
[ ] None of the above

#### Version 2

*Please indicate below whether your biological parents, siblings, or children have ever had cancer*.

Your mother

( ) Yes
( ) No
( ) I’m not sure

Your father

( ) Yes
( ) No
( ) I’m not sure

Your sibling(s)

( ) Yes
( ) No
( ) I’m not sure

Your child(ren)

( ) Yes
( ) No
( ) I’m not sure

How many biological siblings do you have? _____

How many biological children do you have? _____

What type(s) of cancer was your biological mother diagnosed with or treated for? Please select all that apply.

[ ] Bladder cancer
[ ] Brain cancer
[ ] Breast cancer
[ ] Colon/colorectal cancer
[ ] Endometrial or uterine cancer
[ ] Hodgkin’s lymphoma
[ ] Kidney/renal cancer
[ ] Leukemia, any type
[ ] Liver cancer
[ ] Lung cancer
[ ] Myeloma
[ ] Non-Hodgkin’s lymphoma
[ ] Ovarian cancer
[ ] Pancreatic cancer
[ ] Skin cancer
[ ] Stomach cancer
[ ] Thyroid cancer
[ ] Another type of cancer
[ ] I’m not sure

What type(s) of cancer was your biological father diagnosed with or treated for? Please select all that apply.

[ ] Bladder cancer
[ ] Brain cancer
[ ] Colon/colorectal cancer
[ ] Esophageal cancer
[ ] Hodgkin’s lymphoma
[ ] Kidney/renal cancer
[ ] Leukemia, any type
[ ] Liver cancer
[ ] Lung cancer
[ ] Mouth (oral) cancer
[ ] Myeloma
[ ] Non-Hodgkin’s lymphoma
[ ] Pancreatic cancer
[ ] Prostate cancer
[ ] Skin cancer
[ ] Stomach cancer
[ ] Thyroid cancer
[ ] Another type of cancer
[ ] I’m not sure

How many of your biological siblings have ever had cancer? _____

What type(s) of cancer was your biological sibling(s) diagnosed with or treated for? Please select all that apply.

[ ] Bladder cancer
[ ] Brain cancer
[ ] Breast cancer
[ ] Colon/colorectal cancer
[ ] Endometrial or uterine cancer
[ ] Esophageal cancer
[ ] Hodgkin’s lymphoma
[ ] Kidney/renal cancer
[ ] Leukemia, any type
[ ] Liver cancer
[ ] Lung cancer
[ ] Mouth (oral) cancer
[ ] Myeloma
[ ] Non-Hodgkin’s lymphoma
[ ] Ovarian cancer
[ ] Pancreatic cancer
[ ] Prostate cancer
[ ] Stomach cancer
[ ] Skin cancer
[ ] Thyroid cancer
[ ] Another type of cancer
[ ] I’m not sure

How many of your biological children have ever had cancer? _____

What type(s) of cancer was your biological child(ren) diagnosed with or treated for? Please select all that apply.

[ ] Bladder cancer
[ ] Brain cancer
[ ] Breast cancer
[ ] Colon/colorectal cancer
[ ] Endometrial or uterine cancer
[ ] Esophageal cancer
[ ] Hodgkin’s lymphoma
[ ] Kidney/renal cancer
[ ] Leukemia, any type
[ ] Liver cancer
[ ] Lung cancer
[ ] Mouth (oral) cancer
[ ] Myeloma
[ ] Non-Hodgkin’s lymphoma
[ ] Ovarian cancer
[ ] Pancreatic cancer
[ ] Prostate cancer
[ ] Stomach cancer
[ ] Skin cancer
[ ] Thyroid cancer
[ ] Another type of cancer
[ ] I’m not sure

Have any of your biological parents, siblings, or children ever been diagnosed with any of these hereditary cancer syndromes?

[ ] Cowden syndrome
[ ] Familial adenomatous polyposis
[ ] Hereditary breast and ovarian cancer syndrome (HBOC)
[ ] Li-Fraumeni syndrome
[ ] Lynch syndrome (hereditary non-polyposis colorectal cancer syndrome)
[ ] Multiple endocrine neoplasias
[ ] Von Hippel-Lindau Disease
[ ] I’m not sure
[ ] None of the above

## REFERENCES

1. NCCN Clinical Practice Guidelines in Oncology (NCCN Guidelines). Genetic/Familial High-Risk Assessment: Breast and Ovarian. Version 3.2019. Available at: https://www.nccn.org/professionals/physician_gls/pdf/genetics_screening.pdf. Accessed 28 Mar 2019.

2. Committee on Practice Bulletins–Gynecology, Committee on Genetics, Society of Gynecologic Oncology. Practice Bulletin No 182: Hereditary Breast and Ovarian Cancer Syndrome. Obstet Gynecol. 2017;130(3):e110–e126.

3. Rebbeck TR, Kauff ND, Domchek SM. Meta-analysis of risk reduction estimates associated with risk-reducing salpingo-oophorectomy in BRCA1 or BRCA2 mutation carriers. J Natl Cancer Inst. 2009;101(2):80–7.

4. Moyer VA; U.S. Preventive Services Task Force. Risk assessment, genetic counseling, and genetic testing for BRCA-related cancer in women: U.S. Preventive Services Task Force recommendation statement. Ann Intern Med. 2014;160(4):271–81.

5. Gabai-Kapara E, Lahad A, Kaufman B, Friedman E, Segev S, Renbaum P, Beeri R, Gal M, Grinshpun-Cohen J, Djemal K, Mandell JB, Lee MK, Beller U, Catane R, King MC, Levy-Lahad E. Population-based screening for breast and ovarian cancer risk due to BRCA1 and BRCA2. Proc Natl Acad Sci U S A. 2014;111(39):14205–10.

6. Manchanda R, Loggenberg K, Sanderson S, Burnell M, Wardle J, Gessler S, Side L, Balogun N, Desai R, Kumar A, Dorkins H, Wallis Y, Chapman C, Taylor R, Jacobs C, Tomlinson I, McGuire A, Beller U, Menon U, Jacobs I. Population testing for cancer predisposing BRCA1/BRCA2 mutations in the Ashkenazi-Jewish community: a randomized controlled trial. J Natl Cancer Inst. 2015;107(1):379.

7. Metcalfe KA, Poll A, Royer R, Nanda S, Llacuachaqui M, Sun P, Narod SA. A comparison of the detection of BRCA mutation carriers through the provision of Jewish population-based genetic testing compared with clinic-based genetic testing. Br J Cancer. 2013;109(3):777–9.

8. Manickam K, Buchanan AH, Schwartz MLB, Hallquist MLG, Williams JL, Rahm AK, Rocha H, Savatt JM, Evans AE, Butry LM, Lazzeri AL, Lindbuchler DM, Flansburg CN, Leeming R, Vogel VG, Lebo MS, Mason-Suares HM, Hoskinson DC, Abul-Husn NS, Dewey FE, Overton JD4 Reid JG, Baras A, Willard HF, McCormick CZ, Krishnamurthy SB, Hartzel DN, Kost KA, Lavage DR, Sturm AC, Frisbie LR, Person TN, Metpally RP, Giovanni MA, Lowry LE, Leader JB, Ritchie MD, Carey DJ, Justice AE, Kirchner HL, Faucett WA, Williams MS, Ledbetter DH, Murray MF. Exome Sequencing-Based Screening for BRCA1/2 Expected Pathogenic Variants Among Adult Biobank Participants. JAMA Netw Open. 2018;1(5):e182140.

9. Kuchenbaecker KB, Hopper JL, Barnes DR, Phillips KA, Mooij TM, Roos-Blom MJ, Jervis S, van Leeuwen FE, Milne RL, Andrieu N, Goldgar DE, Terry MB, Rookus MA, Easton DF, Antoniou AC; BRCA1 and BRCA2 Cohort Consortium, McGuffog L, Evans DG, Barrowdale D, Frost D, Adlard J, Ong KR, Izatt L, Tischkowitz M, Eeles R, Davidson R, Hodgson S, Ellis S, Nogues C, Lasset C, Stoppa-Lyonnet D, Fricker JP, Faivre L, Berthet P, Hooning MJ, van der Kolk LE, Kets CM, Adank MA, John EM, Chung WK, Andrulis IL, Southey M, Daly MB, Buys SS, Osorio A, Engel C, Kast K, Schmutzler RK, Caldes T, Jakubowska A, Simard J, Friedlander ML, McLachlan SA, Machackova E, Foretova L, Tan YY, Singer CF, Olah E, Gerdes AM, Arver B, Olsson H. Risks of Breast, Ovarian, and Contralateral Breast Cancer for BRCA1 and BRCA2 Mutation Carriers. JAMA. 2017;317(23):2402–2416.

10. King MC, Levy-Lahad E, Lahad A. Population-based screening for BRCA1 and BRCA2: 2014 Lasker Award. JAMA. 2014;312(11):1091–2.

11. Akbari MR, Gojska N, Narod SA. Coming of age in Canada: a study of population-based genetic testing for breast and ovarian cancer. Curr Oncol. 2017;24(5):282–283.

12. Sanchez-Roige S, Fontanillas P, Elson SL, 23 and Me Research Team, Pandit A, Schmidt EM, Foerster JR, Abecasis GR, Gray JC, de Wit H, Davis LK, MacKillop J, Palmer AA. Genome-wide association study of delay discounting in 23,217 adult research participants of European ancestry. Nat Neurosci. 2018;21(1):16–18.

13. Durand, E.Y., Do, C.B., Mountain, J.L., Macpherson, J.M. Ancestry Composition: A Novel, Efficient Pipeline for Ancestry Deconvolution | bioRxiv. Available at: https://www.biorxiv.org/content/10.1101/010512v1. Accessed 28 Mar 2019.

14. Hall MJ, Reid JE, Burbidge LA, Pruss D, Deffenbaugh AM, Frye C, Wenstrup RJ, Ward BE, Scholl TA, Noll WW. BRCA1 and BRCA2 mutations in women of different ethnicities undergoing testing for hereditary breast-ovarian cancer. Cancer. 2009;115(10):2222–33.

15. Manchanda R, Legood R, Burnell M, McGuire A, Raikou M, Loggenberg K, Wardle J, Sanderson S, Gessler S, Side L, Balogun N, Desai R, Kumar A, Dorkins H, Wallis Y, Chapman C, Taylor R, Jacobs C, Tomlinson I, Beller U, Menon U, Jacobs I. Cost-effectiveness of population screening for BRCA mutations in Ashkenazi Jewish women compared with family history-based testing. J Natl Cancer Inst. 2015;107(1):380.

16. Long EF, Ganz PA. Cost-effectiveness of Universal BRCA1/2 Screening: Evidence-Based Decision Making. JAMA Oncol. 2015;1(9):1217–8.

